# Complement activation was not increased in patients with post-acute sequelae after mild SARS-CoV-2 infection: A prospective cohort study

**DOI:** 10.1101/2025.04.26.25326246

**Authors:** Madlene Holmqvist, Dick J. Sjöström, Katherine Carlson, Birgitta Gullstrand, Anders A. Bengtsson, Robin Kahn, Tom E. Mollnes, Per Åkesson, Per H. Nilsson, Fredrik Kahn

## Abstract

Dysregulation of the complement system has been proposed as a pathophysiological mechanism for Post-acute sequelae of SARS-CoV-2 (PASC). We analyzed the complement activation markers C3bc, C3bBbP and TCC in 48 PASC patients, grouped into whether they had a mild (n=38) or severe (n=10) acute SARS-CoV-2 infection, and 80 control subjects. Although the patients with a mild SARS-CoV-2 infection had a trend towards more severe PASC, we could not find any significant differences in complement activation markers between these patients and controls. In conclusion, we could not find convincing evidence of activation of the complement system in PASC patients.

## Background

Post-acute sequelae of SARS-CoV-2 (PASC) is a syndrome that quickly became recognized as a clinical entity during the COVID-19 pandemic. Also known as “long covid”, it occurs in up to one out of eight people with a SARS-CoV-2 infection, affecting an estimated 65 million individuals worldwide^1,2^. Although lacking an exact definition, PASC is often described as persistent or new symptoms that usually last for at least two months after an infection with SARS-CoV-2 and cannot be explained by an alternative diagnosis^3^. The symptoms often include fatigue, dyspnea and cognitive dysfunction, which generally have an impact on everyday activities^3^. PASC does not only affect individuals with an initial severe SARS-CoV-2 infection. In fact, the vast majority of patients with a diagnosis of PASC had a mild acute SARS-CoV-2 infection^5^. Several pathophysiological mechanisms have been proposed including dysregulation of the immune system. T cell alterations, elevated levels of cytokines and autoantibodies are among proposed immune mediated mechanisms^6,7^. In addition, changes in levels of complement factors were detected in this patient group^8,9^. In the present study we evaluated and compared markers of complement activation in both patients with initially mild and severe SARS-CoV-2 infections, who were included at a specialized PASC clinic up to 33 months post-infection.

## Methods

### Study populations

#### PASC patients

Patients ≥18 years old who attended the PASC outpatient clinic at the Department of Infectious Diseases at Skåne University Hospital, a tertiary care hospital serving a population of 1.7 million inhabitants, were eligible for recruitment. Additional inclusion criteria were a test confirming the infection with SARS-CoV-2 (PCR test, antigen test or serology, the latter not as a result of vaccination) and symptoms persisting for at least 12 weeks after the infection. Part of the cohort has previously been described^10^. At inclusion, the patients underwent an interview about their associated symptoms, cognitive tests, sampling of blood and completed questionnaires related to quality of life and symptoms from specific organ systems. The patients were classified in two groups: 1) Patients with a severe acute infection, defined as requiring oxygen therapy during their acute SARS-CoV-2 infection, and 2) Patients with a mild acute infection, defined as not requiring supplemental oxygen during the acute infection. All patients were invited for follow-up visits after 6-12 months, when the same study protocol was repeated, including follow-up plasma sampling.

#### Control subjects

Control subjects ≥18 years old were recruited from participants in other studies with a known history of SARS-CoV-2 infection^11^. The control subjects were completely recovered and reported no persisting symptoms after the infection with SARS-CoV-2 and underwent the same testing procedure as the PASC patients.

### Sampling and complement analyses

Blood samples were collected from patients and controls in 5 ml EDTA-tubes (BD) and centrifuged for 10 min at 2000 x g to obtain plasma. Plasma samples were subsequently stored at −80°C until further use. The complement activation products C3bc, C3bBbP and TCC were measured as markers of activation of the complement system. C3bBbP is a marker specific for activation of the alternative pathway. C3bc is a marker of C3-cleavage and terminal complement complex (TCC) is a marker for complement activation to its very end; both are common to activation of all pathways (classical, lectin and alternative pathways). The assays have been described in detail previously^12^.

### Ethical approval

The present study was approved by the Swedish ethical review authority, (Dnr 2021-03905). All patients and control subjects gave their informed consent to participate in the study.

### Statistics

Statistical analyzes were performed using GraphPad Prism (version 10.3.1 for Mac, GraphPad Software, San Diego, CA), and with R (version 4.4.2) with the packages readxl, MuMin, car (R Core Team (2023). *R: A Language and Environment for Statistical Computing*. R Foundation for Statistical Computing, Vienna, Austria. Medians, interquartile ranges (IQRs), minimum and maximum values were reported when appropriate. Differences between groups were tested with the Mann–Whitney U test, the Wilcoxon signed rank test and Fischer’s exact test as appropriate. P-values less than 0.05 were deemed statistically significant. In the primary analysis, where only the first sample from each person was included, unadjusted p-values were employed. In the secondary analysis where both the first and the second sample was included, p-values were adjusted using the Holm-Bonferroni method to accommodate multiple comparisons. Participants included in the study, but missing blood samples were excluded from the analyses. Missing demographic data were excluded via pairwise deletion.

## Results

### Patient characteristics

Forty-eight patients with PASC and 80 controls subjects were included in this study. The flow chart for inclusion is displayed in Supplementary Figure 1. The patients with long-term symptoms were divided into two groups; 1) severe (oxygen-dependent) and 2) mild (non-oxygen dependent) during the acute infection. The reason for this grouping was the differences in clinical presentation and demographics (Table 1). The pathophysiology for long-term symptoms may also differ between these two groups. Ten patients had a severe acute infection, whereas 38 patients had a mild acute infection. None of the control subjects were hospitalized during the acute infection. Time from acute infection with SARS-CoV-2 to first plasma sampling was a median of 16 months (range 6-26 months) for PASC patients with a severe infection, 15 months (range 9-24 months) for PASC patients with a mild infection, and 10 months (range 5-27 months) for the control group. A second plasma sample was collected in 20 (53%) of the patients with a mild infection and in 6 (60%) of those with a severe infection after a median of 7 months (range 8-10 months) from the first sample.

**Table 1:**
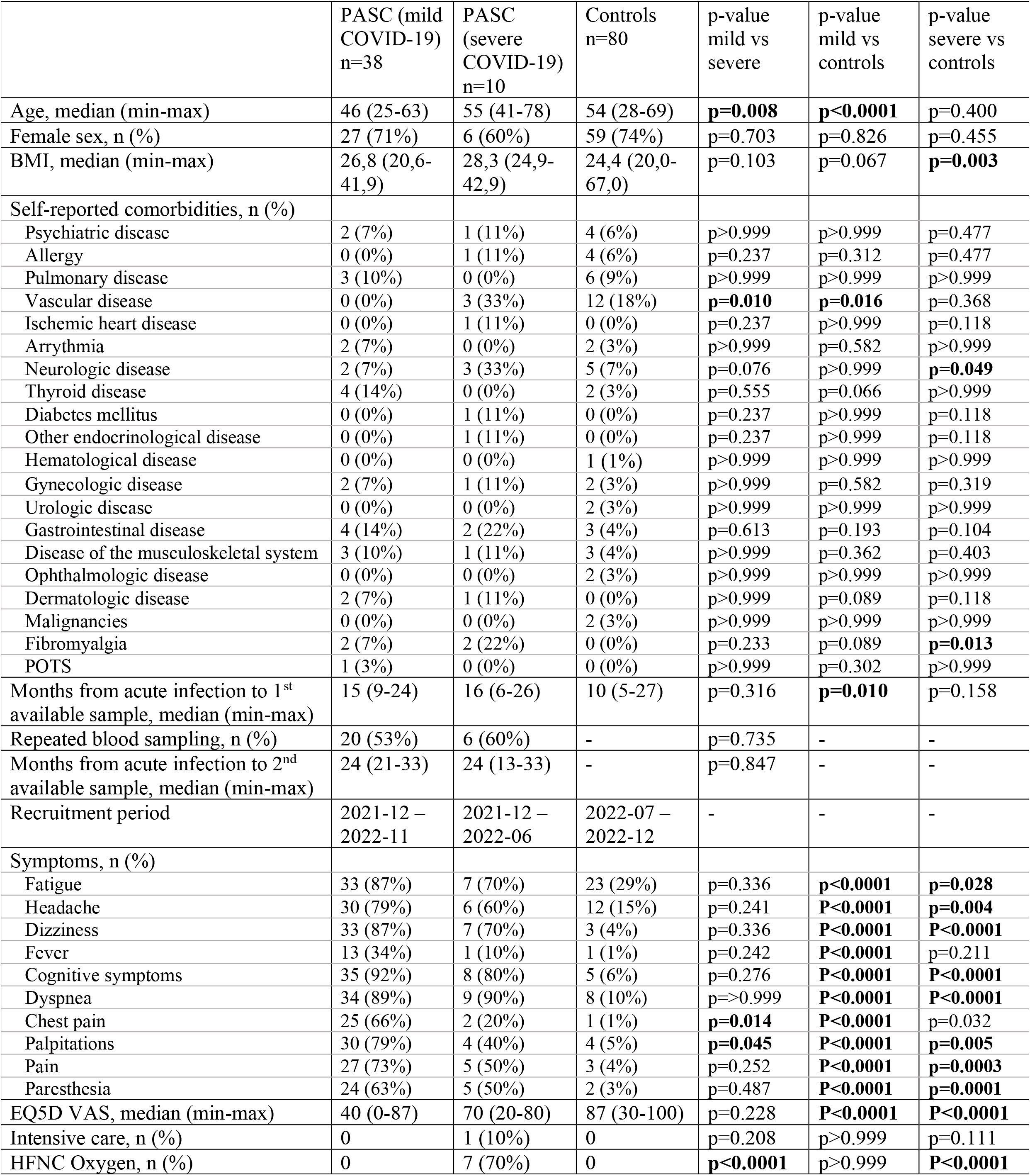
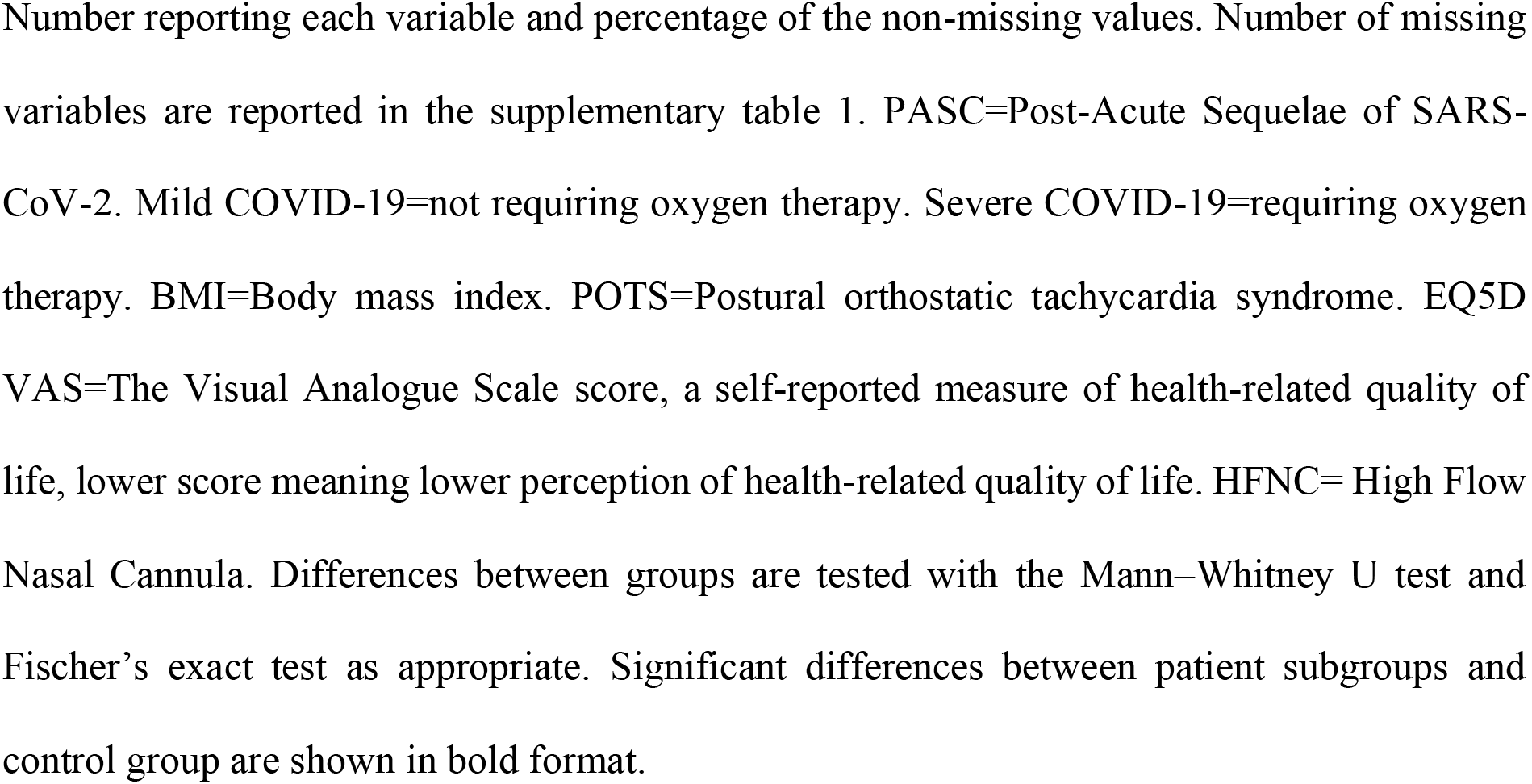
Demographics of the population.

|  | PASC (mild COVID-19)<br>n=38 | PASC (severe COVID-19)<br>n=10 | Controls<br>n=80 | p-value<br>mild vs<br>severe | p-value<br>mild vs<br>controls | p-value<br>severe vs<br>controls |
| --- | --- | --- | --- | --- | --- | --- |
| Age, median (min-max) | 46 (25-63) | 55 (41-78) | 54 (28-69) | **p=0.008** | **p<0.0001** | p=0.400 |
| Female sex, n (%) | 27 (71%) | 6 (60%) | 59 (74%) | p=0.703 | p=0.826 | p=0.455 |
| BMI, median (min-max) | 26,8 (20,6-41,9) | 28,3 (24,9-42,9) | 24,4 (20,0-67,0) | p=0.103 | p=0.067 | **p=0.003** |
| Self-reported comorbidities, n (%) |  |  |  |  |  |  |
| Psychiatric disease | 2 (7%) | 1 (11%) | 4 (6%) | p>0.999 | p>0.999 | p=0.477 |
| Allergy | 0 (0%) | 1 (11%) | 4 (6%) | p=0.237 | p=0.312 | p=0.477 |
| Pulmonary disease | 3 (10%) | 0 (0%) | 6 (9%) | p>0.999 | p>0.999 | p>0.999 |
| Vascular disease | 0 (0%) | 3 (33%) | 12 (18%) | **p=0.010** | **p=0.016** | p=0.368 |
| Ischemic heart disease | 0 (0%) | 1 (11%) | 0 (0%) | p=0.237 | p>0.999 | p=0.118 |
| Arrhythmia | 2 (7%) | 0 (0%) | 2 (3%) | p>0.999 | p=0.582 | p>0.999 |
| Neurologic disease | 2 (7%) | 3 (33%) | 5 (7%) | p=0.076 | p>0.999 | **p=0.049** |
| Thyroid disease | 4 (14%) | 0 (0%) | 2 (3%) | p=0.555 | p=0.066 | p>0.999 |
| Diabetes mellitus | 0 (0%) | 1 (11%) | 0 (0%) | p=0.237 | p>0.999 | p=0.118 |
| Other endocrinological disease | 0 (0%) | 1 (11%) | 0 (0%) | p=0.237 | p>0.999 | p=0.118 |
| Hematological disease | 0 (0%) | 0 (0%) | 1 (1%) | p>0.999 | p>0.999 | p>0.999 |
| Gynecologic disease | 2 (7%) | 1 (11%) | 2 (3%) | p>0.999 | p=0.582 | p=0.319 |
| Urologic disease | 0 (0%) | 0 (0%) | 2 (3%) | p>0.999 | p>0.999 | p>0.999 |
| Gastrointestinal disease | 4 (14%) | 2 (22%) | 3 (4%) | p=0.613 | p=0.193 | p=0.104 |
| Disease of the musculoskeletal system | 3 (10%) | 1 (11%) | 3 (4%) | p>0.999 | p=0.362 | p=0.403 |
| Ophthalmologic disease | 0 (0%) | 0 (0%) | 2 (3%) | p>0.999 | p>0.999 | p>0.999 |
| Dermatologic disease | 2 (7%) | 1 (11%) | 0 (0%) | p>0.999 | p=0.089 | p=0.118 |
| Malignancies | 0 (0%) | 0 (0%) | 2 (3%) | p>0.999 | p>0.999 | p>0.999 |
| Fibromyalgia | 2 (7%) | 2 (22%) | 0 (0%) | p=0.233 | p=0.089 | **p=0.013** |
| POTS | 1 (3%) | 0 (0%) | 0 (0%) | p>0.999 | p=0.302 | p>0.999 |
| Months from acute infection to 1<sup>st</sup> available sample, median (min-max) | 15 (9-24) | 16 (6-26) | 10 (5-27) | p=0.316 | **p=0.010** | p=0.158 |
| Repeated blood sampling, n (%) | 20 (53%) | 6 (60%) | - | p=0.735 | - | - |
| Months from acute infection to 2<sup>nd</sup> available sample, median (min-max) | 24 (21-33) | 24 (13-33) | - | p=0.847 | - | - |
| Recruitment period | 2021-12 – 2022-11 | 2021-12 – 2022-06 | 2022-07 – 2022-12 | - | - | - |
| Symptoms, n (%) |  |  |  |  |  |  |
| Fatigue | 33 (87%) | 7 (70%) | 23 (29%) | p=0.336 | **p<0.0001** | **p=0.028** |
| Headache | 30 (79%) | 6 (60%) | 12 (15%) | p=0.241 | **P<0.0001** | **p=0.004** |
| Dizziness | 33 (87%) | 7 (70%) | 3 (4%) | p=0.336 | **P<0.0001** | **P<0.0001** |
| Fever | 13 (34%) | 1 (10%) | 1 (1%) | p=0.242 | **P<0.0001** | p=0.211 |
| Cognitive symptoms | 35 (92%) | 8 (80%) | 5 (6%) | p=0.276 | **P<0.0001** | **P<0.0001** |
| Dyspnea | 34 (89%) | 9 (90%) | 8 (10%) | p=>0.999 | **P<0.0001** | **P<0.0001** |
| Chest pain | 25 (66%) | 2 (20%) | 1 (1%) | **p=0.014** | **P<0.0001** | p=0.032 |
| Palpitations | 30 (79%) | 4 (40%) | 4 (5%) | **p=0.045** | **P<0.0001** | **p=0.005** |
| Pain | 27 (73%) | 5 (50%) | 3 (4%) | p=0.252 | **P<0.0001** | **p=0.0003** |
| Paresthesia | 24 (63%) | 5 (50%) | 2 (3%) | p=0.487 | **P<0.0001** | **p=0.0001** |
| EQ5D VAS, median (min-max) | 40 (0-87) | 70 (20-80) | 87 (30-100) | p=0.228 | **P<0.0001** | **P<0.0001** |
| Intensive care, n (%) | 0 | 1 (10%) | 0 | p=0.208 | p>0.999 | p=0.111 |
| HFNC Oxygen, n (%) | 0 | 7 (70%) | 0 | **p<0.0001** | p>0.999 | **P<0.0001** |

1 Number reporting each variable and percentage of the non-missing values. Number of missing 2 variables are reported in the supplementary table 1. PASC=Post-Acute Sequelae of SARS- 3 CoV-2. Mild COVID-19=not requiring oxygen therapy. Severe COVID-19=requiring oxygen 4 therapy. BMI=Body mass index. POTS=Postural orthostatic tachycardia syndrome. EQ5D 5 VAS=The Visual Analogue Scale score, a self-reported measure of health-related quality of 6 life, lower score meaning lower perception of health-related quality of life. HFNC= High Flow 7 Nasal Cannula. Differences between groups are tested with the Mann–Whitney U test and 8 Fischer’s exact test as appropriate. Significant differences between patient subgroups and 9 control group are shown in bold format.

Demographic data are displayed in Table 1. In the patient group with a mild SARS-CoV-2 infection, median age was lower compared to the patients with a severe infection (p=0.008) and controls (p<0.0001). The most common PASC-related symptoms in both groups were cognitive impairment, dyspnea, and fatigue. As a measure of health-related quality of life, the participants marked their perception of their current health status between 0-100 with the EQ-5D Visual Analogue Scale (VAS) where 0 represents the worst imaginable health and 100 represents the best imaginable health. PASC patients who had a severe acute COVID-19 disease had a median VAS of 70 (range 20-80), PASC patients who had a mild acute COVID-19 disease had a median VAS of 40 (range 0-87) and the controls had VAS median of 87 (range 30-100). Despite the marked median lower VAS score in the mild group, there was no significant difference between the patients with mild and severe acute COVID-19 disease; presumably due to the large inter-individual differences as well as the small sample size of the severe group.

### Activation of complement factors

Plasma samples were drawn from patients and control subjects both during inclusion as well as patient follow-up visits and were analyzed for the presence of the complement activation products C3bc (common pathway), C3bBbP (alternative pathway) and TCC (terminal pathway). In the first available sample from each individual, no significant differences in C3bc and C3bBbP levels were detected between the two groups of PASC patients and the control group (Figure 1). However, TCC was significantly higher in the patients who had a severe SARS-CoV-2 infection compared to the patients who had a mild acute infection and controls (median 0.70 vs. 0.54 CAU/mL, p=0.041 and median 0.70 vs. 0.52 CAU/mL, p=0.044, respectively), indicating that terminal pathway activation could be associated with severe, but not mild, infection, but not with PASC. C3bc levels in one of the control samples were below the limit of detection and excluded; all other values were within measurable range.

**Figure 1:**
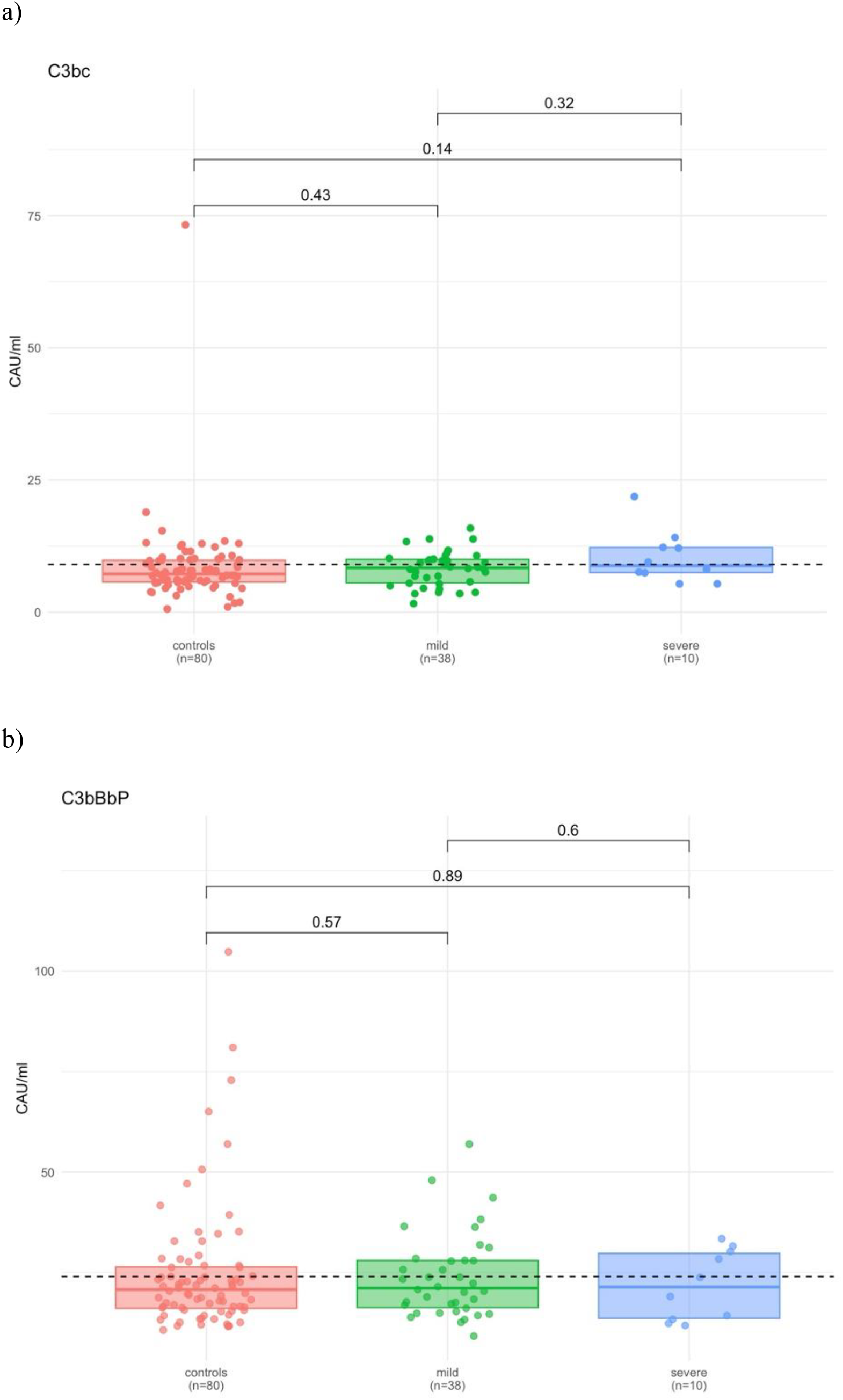
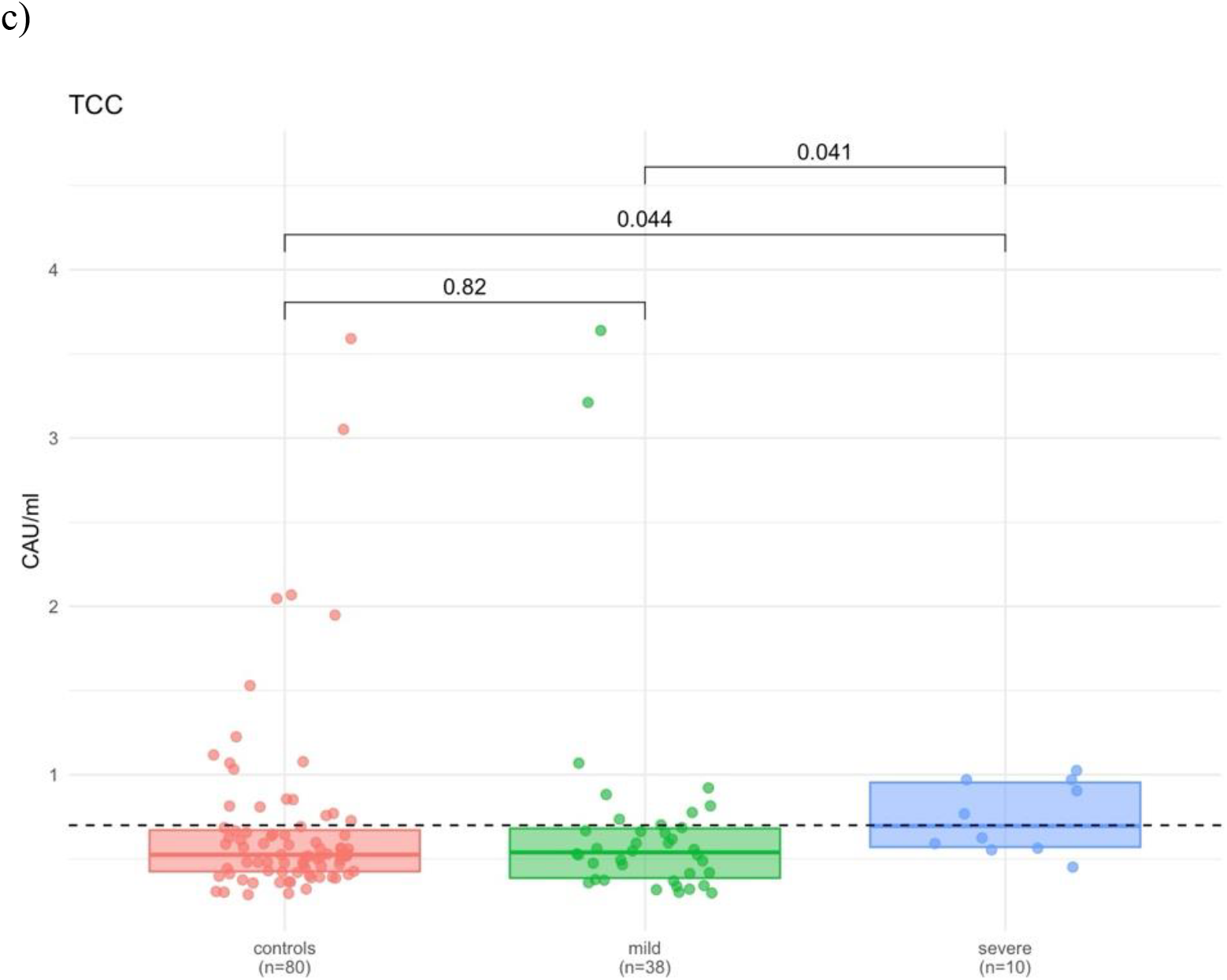
Results of complement activation markers a) C3bc, b) C3bBbP and c) TCC. **a)**Complement activation markers are measured in CAU/mL. Results are demonstrated for the first available sample for PASC patients with a mild and severe acute infection and for control subjects. Differences between groups are tested with the Mann–Whitney U test. A p-value less than 0.05 is considered statistically significant. P-values were unadjusted for multiple comparisons. The dotted line represents upper normal limit as suggested by Bergseth et al^12^.

As a secondary analysis, we analyzed the subgroup of patients with acute mild (n=20) and severe (n=6) SARS-CoV-2 infections who returned for resampling (Supplementary Figure 2). C3bc, C3bBbP and TCC values were not significantly different between the two patient groups compared to controls in neither the first nor the second sample (Supplementary Figure 2). Although there was a statistically significant increase in the activation of C3bBbP between the second and first sample in the mild group, neither sample did significantly differ from controls, suggesting a possible individual variation over time not related to the presence of PASC.

## Discussion

In this study of 48 well-characterized patients with PASC, no significant activation of complement in plasma was detected among persons with an acute mild infection, showing severe PASC. Two recently published studies have shown activation of the complement system in patients with PASC^8,9^. Baillie et al.^8^ studied a cohort of individuals with long-covid and compared them to matched healthy convalescent individuals and detected activation of the classical (C1s-C1INH complex), alternative (Ba, iC3b) and terminal pathway (C5a, TCC). Cervia-Hasler et al.^9^ employed a proteomics approach and found the complement pathway to be dysregulated in patients with PASC compared to controls. They showed increased complement terminal pathway activation in patients with long covid, both during the acute infection and at 6 months follow-up. This imbalance was marked by increased soluble C5bC6 complexes and decreased levels of C7-containing TCC (C5b-9) formations, suggesting increased membrane insertion of TCC (membrane attack complex), contributing to tissue damage in patients with long-covid. Their proteomics data were however recently reanalyzed in a non-peer-reviewed study by Farztdinov et al. who failed to show PASC-dependent complement activation after adjusting the proteomics data for age, body mass index and sex imbalances^13^. We also raise concerns about the sample type in the study by Cervia-Hasler^9^, who employed serum for their complement analysis. Complement activation markers are significantly elevated in serum compared to EDTA-plasma, and analysis of complement activation should thus only be assessed in plasma containing minimum 10 mM EDTA and immediately frozen to −80° C for limiting pre analytical in vitro complement activation^14,15^.

Our cohort differs from the ones in the studies by Cervia-Hasler et al and Baillie et al in three major ways. Firstly, our patients were recruited from a specialized PASC clinic to which they required a referral, which suggests a selection of patients with more severe PASC symptoms. Secondly, in our cohort, we analyzed individuals with an initially mild infection separately from those with an initial severe infection. In the study by Cervia-Hasler et al, these groups were analyzed together, and their cohort comprised of over 70% hospitalized patients and approximately 30% of the cohort had been admitted to the intensive care unit. In the study by Baillie et al. the severity of the acute infection was not described. Notably, we did identify a modest statistically significant elevation of TCC in the group having had an acute severe infection. However, the clinical relevance of this activation remains uncertain. Thus, the difference in the included patients may have given rise to the conflicting results. Thirdly, the time from infection to sampling was in median 16 months in our study, compared to 6 months in the study by Cervia-Hasler et al. In the study by Baillie et al. the median time from infection to sampling is not described. Another difference compared to the study by Cervia-Hasler et al. is that we conducted targeted analyses instead of broad proteomics.

Since no established PASC criteria exist in clinical practice, study design is a challenge. The lack of PASC biomarkers and diagnostic tools present further obstacles leading to notable heterogenic patient cohorts. A strength of this study is that all included patients and control subjects had a documented acute infection with SARS-CoV-2 and underwent thorough interviews on symptom fluctuations. This evaluation was performed by two study physicians to ensure consistent interpretation of patient history. Another strength is separate analyses of patients with mild and severe acute infections, since these groups represents to different entities.

We also acknowledge the limitations of the study, such as the diagnostic uncertainty inherently associated with a diagnosis based on self-reported symptoms as well as relatively small patient cohorts and control groups.

In conclusion, we could not find convincing evidence of activation of the complement system in patients with PASC due to an acute mild SARS-CoV-2 infection, the most frequent clinical scenario preceding this condition^5^. Thus, our data do not support the theory of complement activation as a key pathophysiological mechanism.

## Data Availability

All data produced in the present study are available upon reasonable request to the authors

## Acknowledgments

We acknowledge the work of our dedicated research nurses Alejandra Castilla, Tine Søraas and Sara Hansson. ChatGPT-4 was utilized for coding assistance and debugging, and for language improvement.

## Author contributions

FK conceived the study, with contributions from RK, AB, BG, PÅ, PN and TEM. MH and KC recruited patients and collected clinical data. DS and PN analyzed the complement assays. MH and FK conducted the statistical analyses. MH drafted the manuscript with assistance from PÅ and FK. All authors contributed with interpretation of results, critically revised the manuscript and approved the final version for submission.

## Supplementary material

**Supplementary figure 1:**
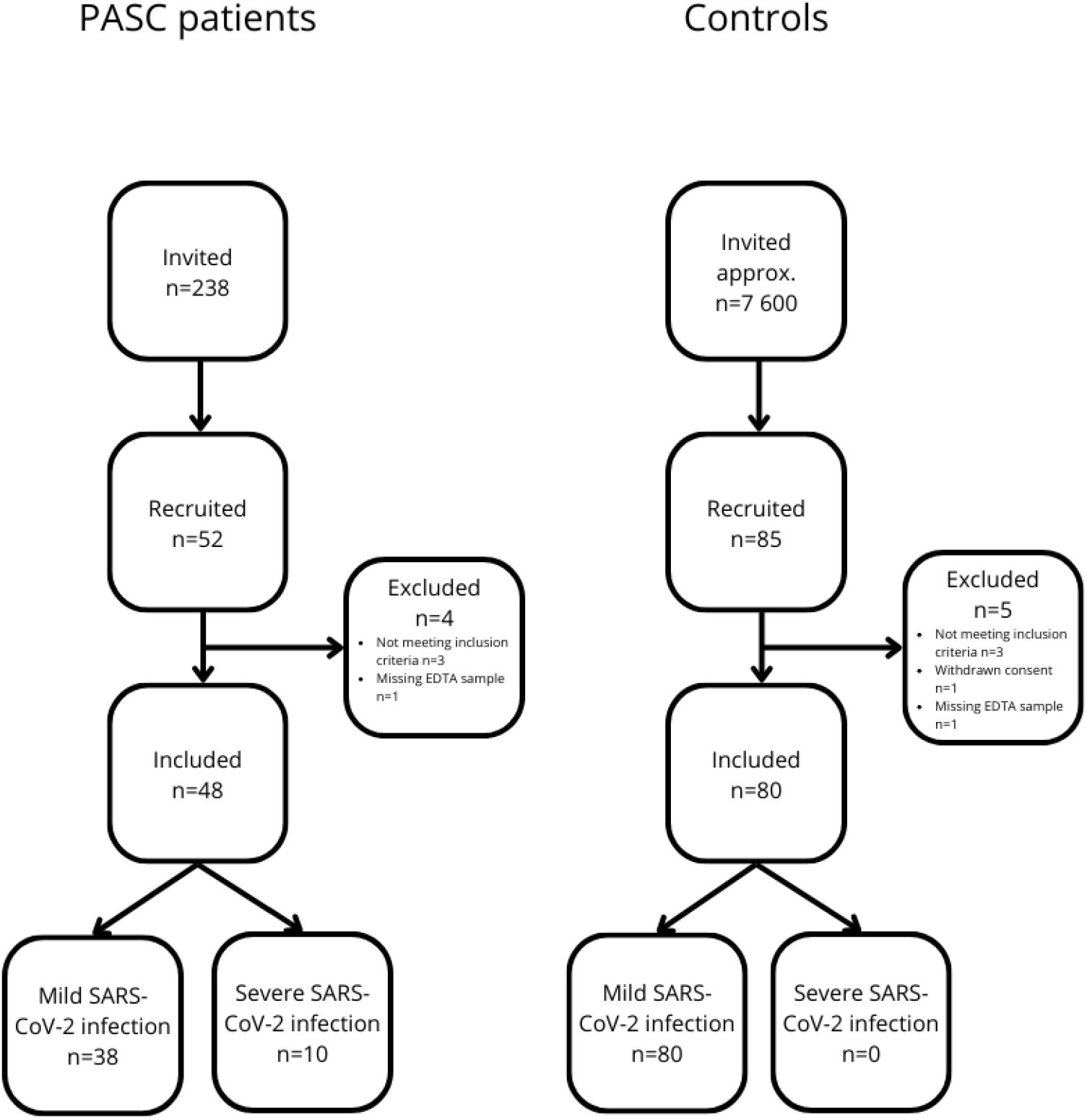
Flow chart for inclusion of PASC patients and control subjects.

**Supplementary figure 2:**
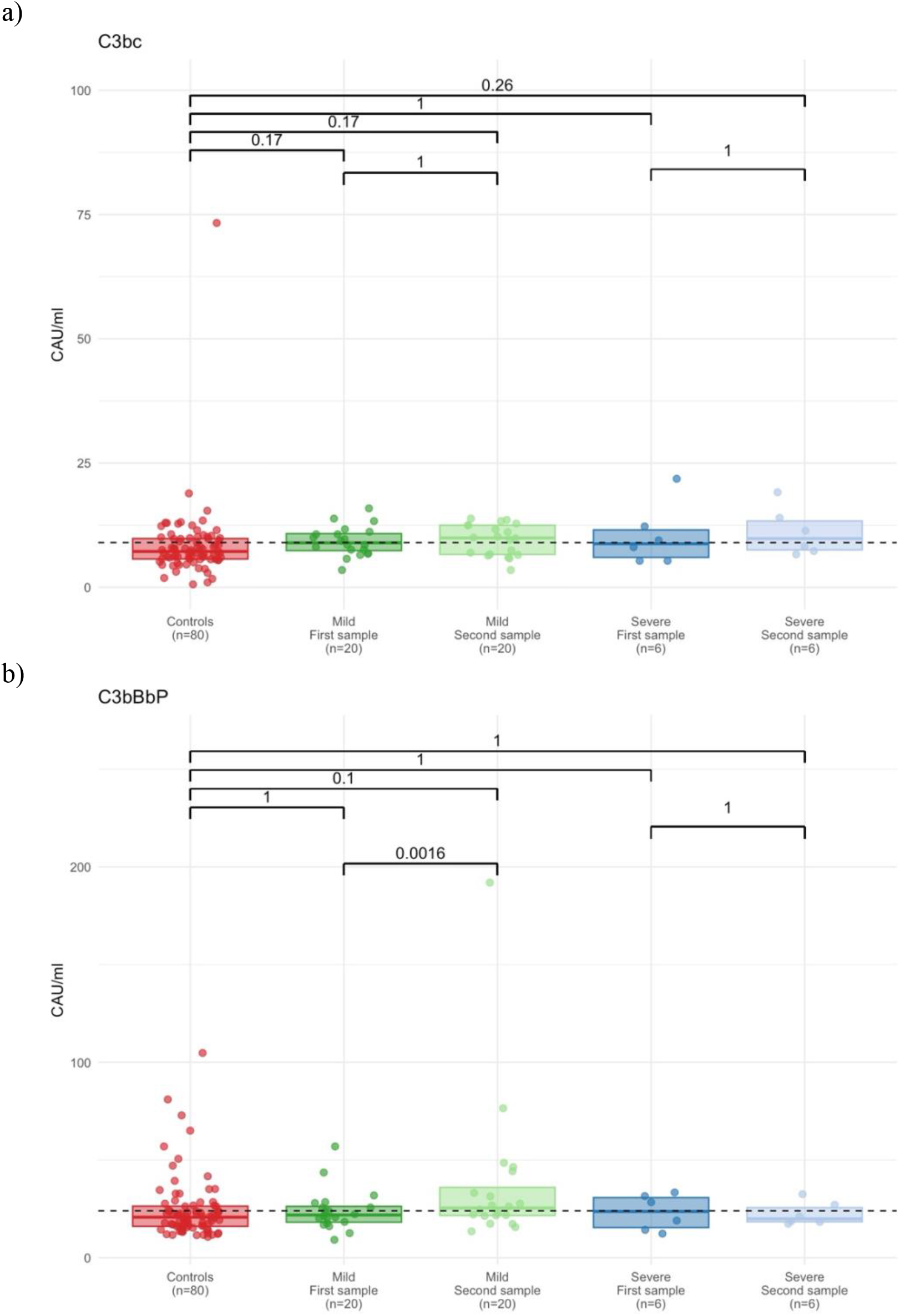
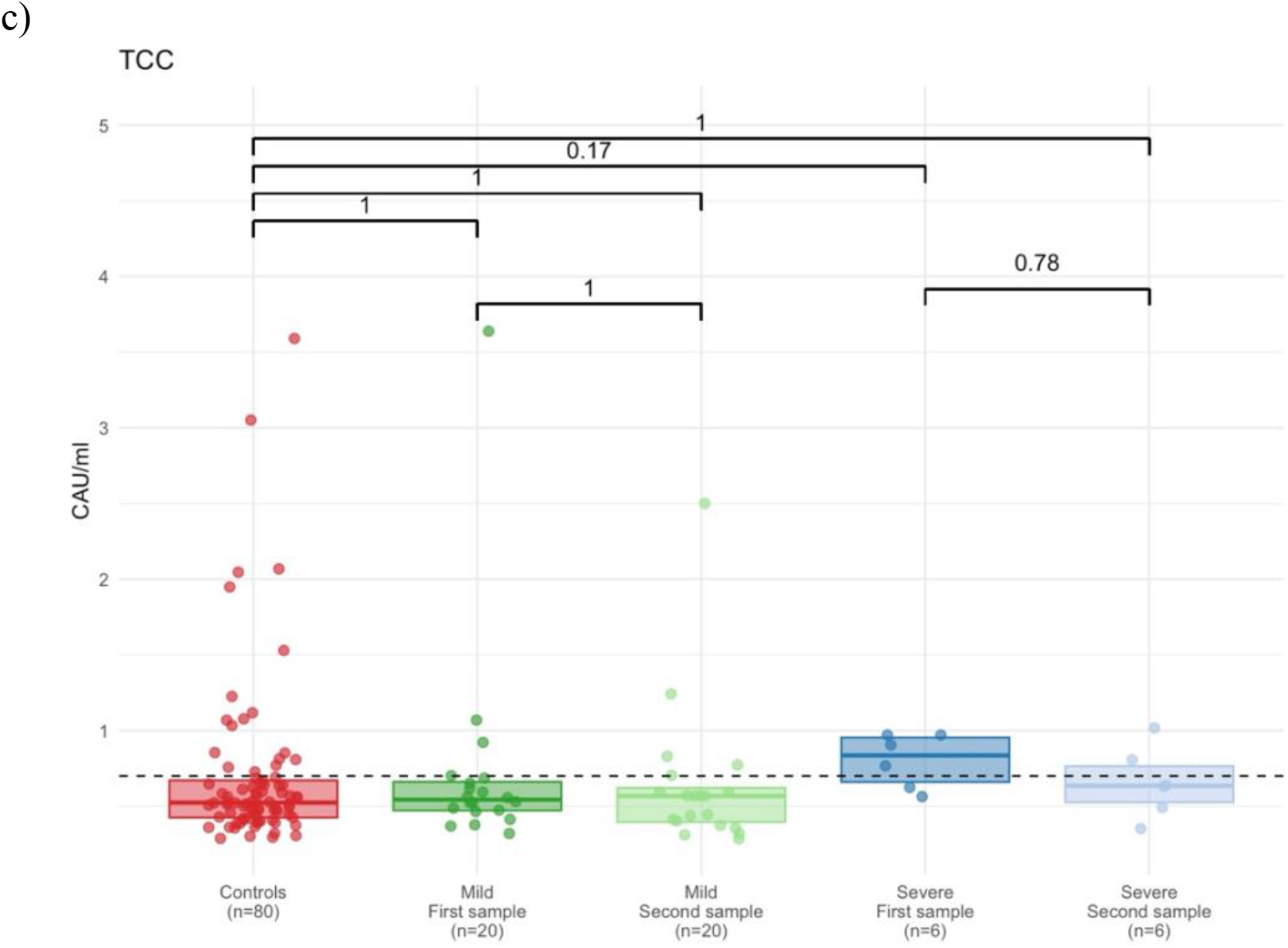
Secondary analysis of repeated measurement of complement activation markers a) C3bc, b) C3bBbP and c) TCC. Complement activation markers are measured in CAU/mL. Differences between group medians are tested with the Mann–Whitney U test and differences between paired measurements are tested with the Wilcoxon signed rank test. A p-value less than 0.05 is considered statistically significant. P-values are adjusted within every complement activation marker respectively, using the Holm-Bonferroni method to accommodate multiple comparisons. The dotted line represents the upper normal limit as suggested by Bergseth et al^12^.

**Supplementary table 1:** Missing data from table 1.

|  | PASC (mild<br>COVID-19)<br>n=38 | PASC<br>(severe<br>COVID-19)<br>n=10 | Controls<br>n=80 | 2<br>3<br>4 |
| --- | --- | --- | --- | --- |
| Age, n (%) | 0 | 0 | 0 |  |
| Female sex, n (%) | 0 | 0 | 0 | 5 |
| BMI, n (%) | 9 (24%) | 2 (20%) | 10 (8%) |  |
| Self-reported comorbidities, n (%) | 9 (24%) | 1 (10%) | 13 (16%) | 6 |
| Months since acute infection to 1<sup>st</sup><br>available sample, n (%) | 0 | 0 | 0 | 7 |
| Months since acute infection to 2<sup>nd</sup><br>available sample, n (%) | 0 | 0 | - | 8 |
| Symptoms, n (%) | 0-1 (0-3%) | 0 | 0-1 (0-1%) |  |
| EQ5D VAS, n (%) | 9 (24%) | 1 (10%) | 13 (16%) | 9 |
| Intensive care, n (%) | 0 | 0 | 0 |  |
| HFNC Oxygen, n (%) | 0 | 0 | 0 | 10 |

Number and proportion of missing values from table 1.

